# Translation, adaption and validation of HIV/AIDS stigma and discrimination scale for university students in the general population of China

**DOI:** 10.64898/2026.03.06.26347320

**Authors:** Xiaohan Wang, Jingjing Zhao, Runqing Liu, Zhe Wu, Xiang Chen, Zhengyang Pan

**Author notes:** These authors contributed equally to this work (Xiaohan Wang, Jingjing Zhao, Runqing Liu, Zhe Wu). **Correspondence to:** Zhengyang Pan, The First School of Clinical Medicine, Anhui Medical University, Hefei, China. **Conflicts of interest:** None.

## Abstract

**Background:** Stigma – a procedures with label, stereotype, prejudice, status loss, and discrimination –has long played a role in the spread of HIV since the beginning of the epidemic. However, few researchers conducted on the HIV-related stigma and discrimination for general population in China. Consequently, we introduced translated and adapted the English version of HIV/AIDS Stigma and Discrimination Scale applied for undergraduates in China.

**Objective:** This study aimed to adapt the HIV/AIDS Public Stigma and Discrimination Scale (HPSDS) in China and to investigate its psychometric properties (e.g., reliability and validity).

**Methods:** Using translation, back-translation, quality evaluation, cross-cultural adaption and pre-assessment, a Chinese draft version of the HPSDS was obtained. From April 2022 to July 2022, the scale was distributed to179 universities and colleges and 2,333 college students filled out the translated and adapted questionnaires. Finally, we collected 1,604 valid questionnaires. The results were recruited to assess the psychometric characteristics of the CV-HPSDS.

**Result:** The CV-PHSDS consists of 3 dimensions and 14 items with Cronbach’s alpha coefficient, McDonald’s omega coefficient and test-retest reliability of the scale are 0.869, 0.883 and 0.857 respectively, manifesting good internal consistency and stability. To construct validity of adapted scale, an exploratory factor analysis was conducted with the cumulative variance contribution rate of 76.6% was obtained. For confirmatory factor analysis, the CFI, GFI, TLI, and IFI showed excellent fitness to the structure, with fitness indices of 0.972, 0.949, 0.965, and 0.972, respectively. Finally, a valid and reliable instrument to measure the HIV/AIDS stigma and discrimination level is obtained.

**Conclusion:** The translated and adapted version of HPSDS shows to be a reliable and valid instrument for assessing stigma and discrimination level for undergraduates.

## Introduction

HIV/AIDS contributed to majority of burden for sexual healthcare disease related worldwide, while antiretroviral therapy have turned HIV/AIDS into a chronic but manageable progression. For people living with HIV (PLHIV)/AIDS, there were profound and lasting impact on life-quality among whom patients persisted regular therapy, and it remained a more influential public issues throughout life. There were 39.9 million PLHIV with 630,000 deaths in 2023 globally, among which 6.7 million of PLHIV dwelled in the Asia-Pacific region [1]. Worse condition occurred in young population. It was estimated that DALYs of 426.4 and incidence of 39.7 per 100,000 population within young adults [2]. However, PLHIV often fall short in unfair treatment—labelled, stereotyped, and discrimination, for decades that contributed to worse consequences than disease itself [3][4][5].

Substantial efforts have been made to improve high-quality life for PLHIV and more friendly condition for HIV/AIDS community. In the context of achieving the ambitious “95-95-95-95 target” (95% combination prevention, 95% detection, 95% treatment, 95% viral suppression) by 2030[6], the stigma and discrimination have been identified as the most important barrier to effective prevention and efficient response in China and worldwide[3][7][8]. Removing HIV-related stigma is crucial for achieving global health goals, and assessing such stigma constitutes. Especially for public stigma, also called interpersonal stigma, it is defined as negative attitudes and beliefs that motivate individuals to keep social distance and avoid interaction with stigmatized groups[9]. In addition, states of the general population attitude, especially young adolescent students, towards PLHIV have positive influence on decreasing stigma and discrimination (SAD), implementing people-centered HIV services and restraining further transmission[10][11].

The potential aimed population of eliminating stigma and discrimination may extend beyond perceived stigma, for whom PLHIV deemed that they were insulted, rather than construct de-/non-stigmatized ecosystem for HIV/AIDS. In a previous study by XX et al., it showed that PLHIV perceived SAD from families, communities and healthcare settings[12], the existing research is narrow to understand what the general population actually considers about how to treat their HIV-positive neighbors. Notably, HIV-positive women perceive a higher level of stigma than the level of stigma expressed by people in the community[13]. Although previous research has developed several scales or questionnaires, their limitations lie in the fact that the assessment by the scale is not comprehensive[14][15]. Meanwhile, other antecedent scales have been designed for specific professions or employed samples with limited sources[5][16][17]. This has led to only a few scales being available and comprehensive for the general population to evaluate and assess the level of SAD. Consequently, the main objective of this study is to translate, adapt and assess the Chinese version HIV/AIDS public stigma and discrimination scale (CV-HPSDS) for general population. The original version of the scale was developed and underwent psychometric testing in Iran[16]. We hope that the short-version CV-HPSDS will be instrumental in evaluating the level of SAD with large-scale application and will be helpful in performing future interventions.

## Methods

### Study Design and Participants

The present study was designed to undergo translation, back-translation, cultural adaptation, and psychometric assessment. The cross-translation process was carried out using adapted Brislin’s model, which involves five steps[18].

### Translation and Cross-adaption

The research team established contact with Professor Mahlagha Dehghan and received permission to translate the original version of HIV/AIDS Public Stigma and Discrimination Scale from the perspective of the general population (OV-HPSDS). The team was guided and supervised by Professor Dehghan in their subsequent work. The concept of SAD has suffered from “conceptual inflation”, where SAD has been mixed and confused[19][20]. In this study, stigma refers to the experience of being devalued, stereotyped, and labeled, and discrimination deals with the act of being rejected and disregarded[21].

To ensure accurate translation, we formed two independent groups. The first group comprised two English teachers with 15 years of experience in English, both of whom work at Anhui Medical University. They translated the original scale independently and discussed each item thoroughly. Both of them have experience in translation, overseas academic experience, and knowledge of healthcare terminology. They provided us with the Initial Translation-1 (TL-1). The second group comprised two bilingual individuals who were not familiar with the questionnaire concepts and did not have a medical background. They translated the items independently and provided us with the Initial Translation-2 (TL-2). By using two different sets of bilingual translators with diverse backgrounds, we ensured that the content was culturally appropriate for China and achieved conceptual equivalence.

The translations (TL-1) and (TL-2) were carefully reviewed, discussed, and synthesized during a consensus meeting between the translators and researchers. This stage led to the development of the pre-final translation (PTL-1), which was achieved after the researchers reconciled and compared any ambiguities to the original version. The items in the PTL-1 were thoroughly discussed and a consensus was reached, ensuring that the translated version accurately reflected the intended meanings.

The expert consultancy was composed of all the translators and researchers, and they reviewed the pre-final version of the CV-HPSDS based on the PTL-1. The translated version was then sent to experts who have extensive experience in public health and HIV care. These experts identified that some items were insufficient or inappropriate. In particular, they suggested that certain items containing illegal behaviors that infringe the rights of PLHIV should be omitted from the pre-final version. Finally, our team formed Adapted-PTL-1 (APTL-1).

Two bilinguals with overseas living experience and psychology abilities were selected to carry out the back-translation process based on the PTL-1. The translators did not have a medical background and were not familiar with the original instrument. They independently produced two new back-translation versions, BTL-1 and BTL-2. After completing their work, the experts discussed and synthesized the two back-translation versions to create the final version back-translation (FBTL). The researchers ensured the semantic, idiomatic and conceptual equivalence of the translated version. To assess the equivalence of CV-HPSDS, we sent the final version of the scale to the original author along with an explanation of the rationale behind the adjustments made. The primary author confirmed that the FBTL reflected the keynote of the OV-HPSDS.

### Per-experiment test

In the preliminary experiment, 117 students were recruited to evaluate the translated version of the questionnaire. The participants were asked to provide feedback on the intelligibility, appearance, clarity, and wording of the items. The reliability of the questionnaire was estimated based on the results of the preliminary test. The internal consistency of the questionnaire was found to be high, with Cronbach’s alpha coefficient and split-half coefficient values of 0.851, 0.879, and 0.779, respectively. Exploratory factor analysis was used to evaluate the structure of the questionnaire, resulting in a 3-factor model that was deemed suitable for the questionnaire (KMO=0.835, Bartlett’s chi-square=1705.631, P<0.001). Although this model differed from the original 4-factor model, it had a better cumulative explained rate of 68.307%. To ensure better construct validity, only items with factor loadings above 0.4 were retained in the same dimension, except for item 8. The item-total correlation ranged from 0.262 to 0.751 (P<0.001), and the item-dimension correlation ranged from 0.499 to 0.927 (P<0.01). Overall, these results demonstrate that the translated version of the questionnaire is acceptable.

The cultural adjustments made to the items in PTL-2 were found to be suitable for the Chinese context after the expert review and pilot test. For example, the expression order of some items was adjusted to fit the Chinese custom of modifying adverbial modifiers and clauses. Additionally, item 8 was deleted from the scale as it was deemed inappropriate for the Chinese culture and did not fit the scale structure. The expert reviewers pointed out that blood relationships are highly valued in traditional Chinese virtues, and families do not abandon their members. After passing the pilot test, the 17 items in PTL-2 were included in the formal investigation to evaluate the psychometric parameters.

### Formal survey

From April 2022 to July 2022, 2195 college students from 179 universities in China were recruited for the formal survey and 1456 valid questionnaires were selected out, giving a valid response rate of 71.66%. 31 students were randomly invited and finished the test-retest survey with a 2-week interval. Additionally, the data were collected through a web-based investigation platform (https://www.wjx.cn/). The survey followed strict anonymous principles and informed consent was obtained from all subjects.

## Instrument

### Demographic Information Questionnaire

Following a thorough literature review, the research team constructed a questionnaire that adhered to the scale, aimed at collecting fundamental demographic characteristics such as gender, local institution, education level of parents, grades, and whether the subjects were majoring in medical fields or working in medicine-related occupations. Additionally, the questionnaire aimed to assess the subjects’ basic knowledge regarding HIV condition in the context of their social background. For instance, within the group of adults who have worked, questions such as “If you are aware of any friends or relatives with HIV/AIDS” and “If you have undergone HIV/AIDS testing” were included.

### Chinese version HIV/AIDS Public Stigma and Discrimination Scale

The Iranian scale initially comprised 18 items that were distributed across 4 dimensions. However, following a pre-experiment survey, the CV-HPSDS was reduced to 17 items and 3 dimensions, namely social disease perspective (6 items), social support (4 items), and patient social discrimination (7 items) for the formal survey. All items were rated on a Likert-5 scale, ranging from 1 to 5 (extremely disagree to extremely agree). Higher scores indicated higher levels of stigmatization held by the subjects towards PLHIV.

### Date Collection

The distribution of questionnaires to an adequate sample size was carried out by trained researchers who had participated in a consensus meeting aimed at harmonizing the items in the scale. Thus, the researchers were well-versed with the meaning and conceptualization of the items. An online survey was administered through a web-based investigation platform (https://www.wjx.cn/).

### Statistical Analysis

The collected data were inputted into SPSS 26.0 and AMOS 23.0 for statistical analysis in order to determine the reliability and validity of the scale. Descriptive statistics were used to summarize the data and reported as mean± standard deviation (SD). All statistical tests were two-tailed and had an alpha of 5% (p<0.05), which was considered statistically significant. Any significant findings were further explored to provide a more in-depth understanding.

### Item analysis

The present study utilized item analysis to evaluate the discrimination and homogeneity of the items. Specifically, the Critical Ratio method was employed, which involved calculating the total score and selecting the top and bottom 27% of the score distribution for high and low groups, respectively. A sample t-test was then conducted to compare the mean scores of the two groups, and items that showed statistically significant differences (p<0.05) were retained. The question total correlation method was also employed, which examined the correlation between each item and the total score of the scale. Items that showed significant correlations (p<0.05) or a total correlation coefficient of less than 0 were retained. Additionally, the corrected item-total correlation (CICT) was calculated to assess the correlation between each item and the total scale. Items with a CICT of 0.5 or higher were considered to demonstrate acceptable internal consistency and stability. Furthermore, Cronbach’s alpha coefficient was employed to evaluate the internal consistency of the scale, with items being removed if they resulted in a lower coefficient. Our study suggests that lower Cronbach’s coefficients indicate the need to improve the internal consistency of the scale.

### Exploratory factor analysis

Given the cultural differences and the need for cross-adaptation for better application to the Chinese version, an exploratory factor analysis (EFA) was implemented to synthesize a large number of items and group them into dimensions to determine the factor structures. In addition, the Kaiser-Meyer-Olkin (KMO) measure and the Bartlett’s test of sphericity were used to test the factorability of the scale. If the KMO is greater than 0.7, the model is considered suitable for analysis. EFA is a statistical method used to assess the construct validity of a scale. It extracts underlying factors and applies varimax rotation to simplify the scale’s structure, making the factors easier to interpret.

### Confirmatory factor analysis

The validity of the scale was confirmed through the use of a theoretical model by employing AMOS26.0, thereby ascertaining the model in EFA. The Root Mean Square Error of Approximation (RMSEA), χ2/degree of freedom, comparative fit index (CFI), goodness-of-fit index (GFI), Tucker-Lewis index (TLI), and incremental fit index (IFI) were used to measure whether the model fits for the structure of the scale. The higher the CFI, GFI, TLI, and IFI, the better the model for the scale’s structure. Furthermore, a result of less than 0.1 for the RMSEA indicates good fitness for the model.

### Internal Consistency

The reliability of the scale was assessed using two methods, namely test-retest research and the Cronbach alpha coefficient which was employed to examine the internal consistency of the scale. A Cronbach’s alpha coefficient value > 0.7 was considered suitable, indicating an acceptable psychometric tool for further research. Moreover, McDonald’s omega coefficient was computed to evaluate the reliability of the scale, as it is a more suitable parameter for multidimensional tests compared to the Cronbach alpha coefficient. An omega coefficient >0.8 indicated an acceptable reliability of the scale. Additionally, the composite reliability (CR) was used as an indicator of the shared variance of the observed variables and the total amount of true score variance related to the total, providing a more accurate method for assessing reliability with a threshold > 0.6.

### Known-groups Validity

The known-group test serves as a crucial step towards reinforcing the construct validity of the instrument. To execute this test, the sample was divided into two groups to evaluate the differences in the hypotheses of the scale. The study hypothesized that female subjects, who were educated in university, majoring in medical fields and residing in urban areas, would have lower scores on the total scale, while the other scores would be lower.

### Convergent validity and discriminant validity

The assessment of convergent validity and discriminant validity was carried out using the average variance extracted (AVE). The former measures the extent to which constructs that should theoretically be related are indeed related. An AVE value exceeding 0.5 indicates strong construct validity. Meanwhile, the latter assesses whether the constructs in the model exhibit low intercorrelations. Furthermore, acceptable discriminant validity is demonstrated if the square root of AVE exceeds the correlations in subscales.

## Results

### Demographic Characteristics

A total of 2195 questionnaires were collected, and valid data were obtained from 1456 respondents, resulting in a valid response rate of 71.66%. Approximately one-quarter of the participants were students in the first grade(27.47%). Of the total sample, 894 (61.40%) resided in urban areas, and the remaining 562 (38.60%) lived in rural areas. Regarding gender, 776 (53.30%) respondents were male and 680 (46.70%) were female. Few participants had been tested for HIV (9.55%) or had contact with PLHIV (3.02%). Detailed characteristics were shown in Table 1.

**Table 1.** Demographic characteristics of the subjects (N=1456)

| Independent variable | Sample (n=1456) |  |
| --- | --- | --- |
|  | n | % |
| **X1: Gender** (female) | 680 | 46.70 |
| male | 776 | 53.30 |
| **X2: Residence** (urban) | 894 | 61.40 |
| Rural | 562 | 38.60 |
| **X3: Major:** (Medical) | 848 | 58.24 |
| Non-medical | 608 | 41.76 |
| **X4: Grades** (Freshman) | 400 | 27.47 |
| Non-freshman | 1056 | 72.53 |
| **X5: School** (University) | 1310 | 89.97 |
| College | 146 | 10.03 |
| **X6: Father's education** |  |  |
| (Below the primary school) | 83 | 5.70 |
| Primary school/Junior high school | 521 | 35.78 |
| Senior high school/vocational high school | 437 | 30.01 |
| College/technical secondary school | 371 | 25.48 |
| Bachelor degree or above | 44 | 3.02 |
| **X7: Mother's education** |  |  |
| (Below the primary school) | 143 | 9.82 |
| Primary school/Junior high school | 471 | 32.35 |
| Senior high school/vocational high school | 367 | 25.21 |
| College/technical secondary school/ | 426 | 29.26 |
| Bachelor degree or above | 49 | 3.37 |
| **X8: Whether to conduct the test of HIV/AIDS** (No) | 1317 | 90.45 |
| (Yes) | 139 | 9.55 |
| **X4: whether to make acquaintance with people living with HIV/AIDS or relatives infect with HIV/AIDS** (No) | 1412 | 96.98 |
| (Yes) | 44 | 3.02 |

### Items analysis

The results of the critical ratio test demonstrated that all items were significantly different in the high and low groups (p<0.01). Furthermore, the correlation coefficients of the items-total correlation ranged from 0.322 to 0.798, with all reaching significant differences (p<0.001). Analysis of the Cronbach’s coefficient and CICT revealed that four items were unsuitable for the scale. Although removing these items would improve the Cronbach’s coefficient and correlation between the total and items, we chose to retain them because they belonged to the same subscale, “social support”, and were related to discrimination behavior in various domains of life. The “social support” subscale was also an important factor for our research team. (Table 2)

**Table 2.**
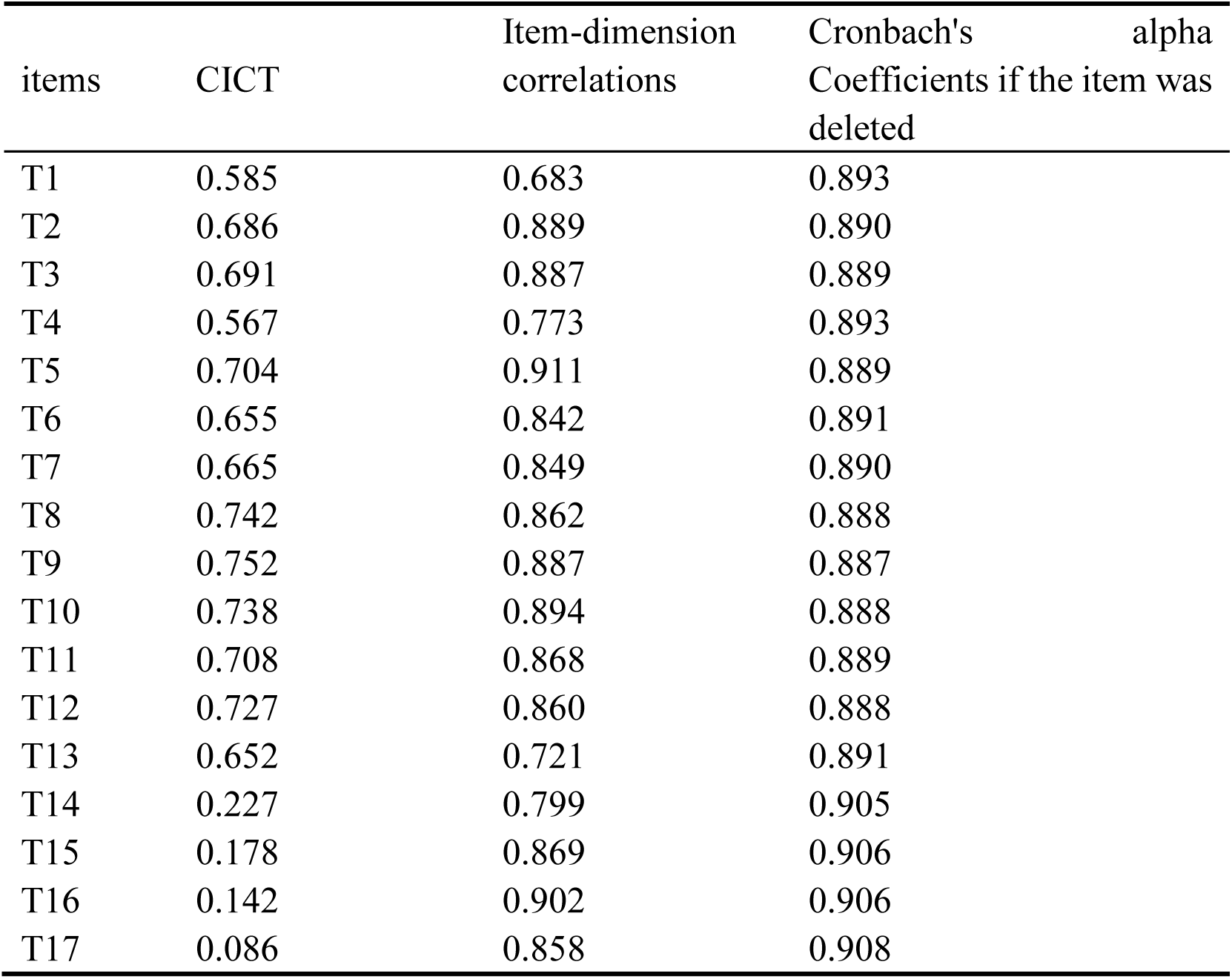
The internal consistency of items and the corrected item-total correlation (n=1,456)

### Exploratory factor analysis

The results of the exploratory factor analysis (EFA) of the scale indicated a Kaiser-Meyer-Olkin (KMO) value of 0.929, and the Bartlett’s test of sphericity was found to be statistically significant (χ2=19961,026, p<0.001). Therefore, we conducted factor analysis using Principal Axis Factoring (PAF) with varimax rotation. Based on the eigenvalues exceeding than 1, we identified 3 factors according to the Scree Plot. The cumulative variance contribution rate was found to be 73.313%.

It is noteworthy that in accordance with the principle of excluding cross-factor items, PSD 1 and PSD 13 were eliminated from further analysis. The remaining 15 items were subjected to EFA once again. A Scree Plot was employed to assess the eigenvalues, which were found to be 6.856, 3.113 and 1.520. A KMO value of 0.921 and significant Bartlett’s test (χ2=18786.223, p<0.001) confirmed the suitability of the data for factor analysis. The cumulative variance contribution rate of 76.597% was obtained. (Table 3)

**Table 3.**
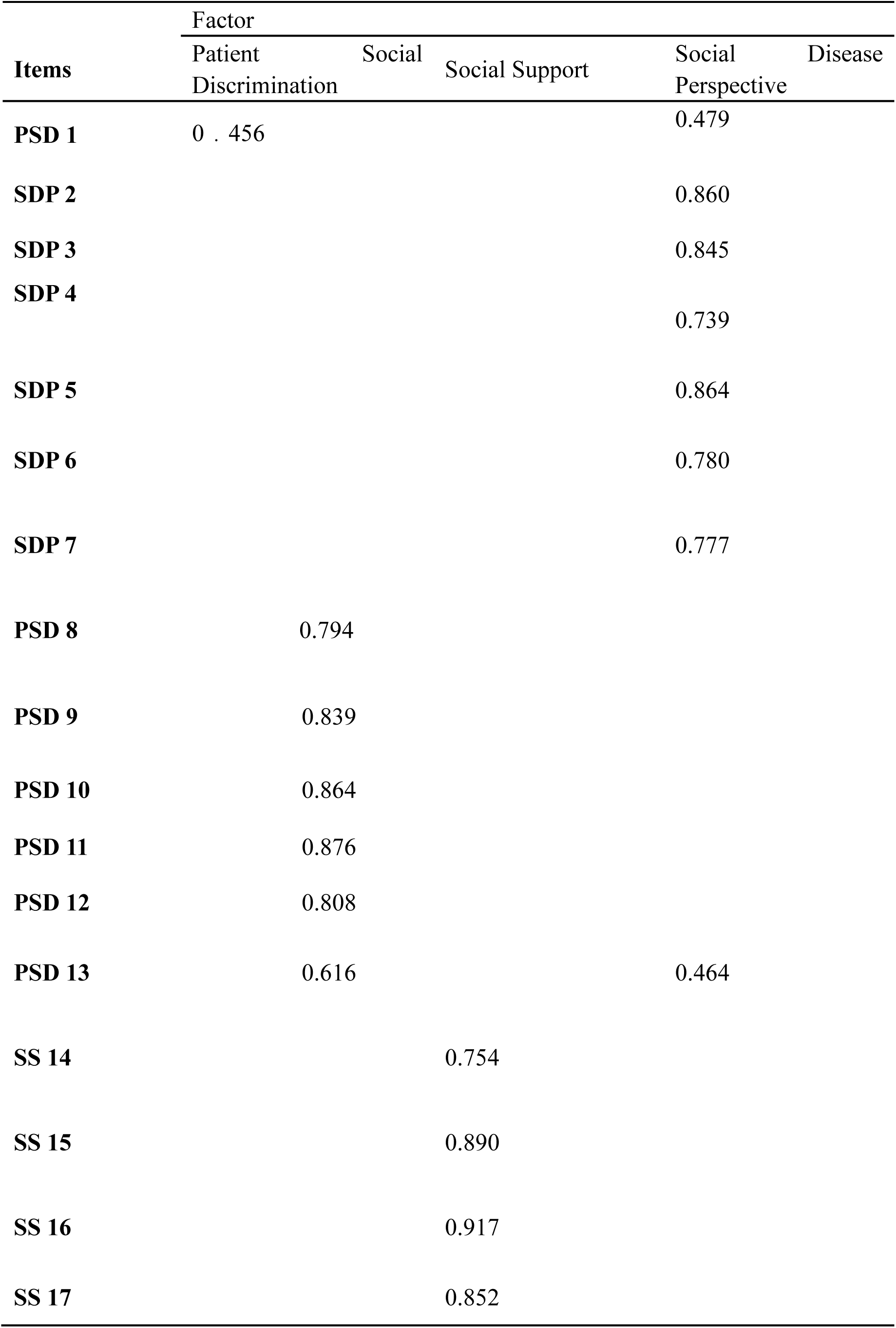
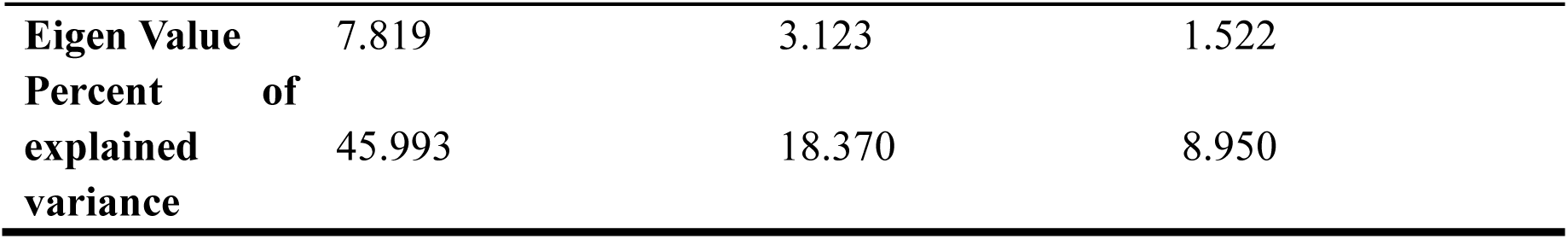
Exploratory Factor Analysis of HIV/AIDS Public Stigma and Discrimination Scale.

### Confirmatory factor analysis

After identifying a 3-factor model, we conducted CFA to confirm the factor structure derived from EFA. However, the results indicated that the associated level of chi-square failed to meet the required standard, rendering the model unacceptable. As suggested by the parameters in the AMOS analysis, we removed item 11 and included the remaining 14 items in the 3-factor model. (Figure 1) All of the fit indices reached suitable levels. The RMSEA and Standardized Root Mean Square Residual (SRMR) achieved acceptable levels, while the CFI, GFI, TLI, and IFI showed excellent fitness to the structure, with fit indices of 0.972, 0.949, 0.965, and 0.972, respectively. (Table 4)

**Figure 1.**
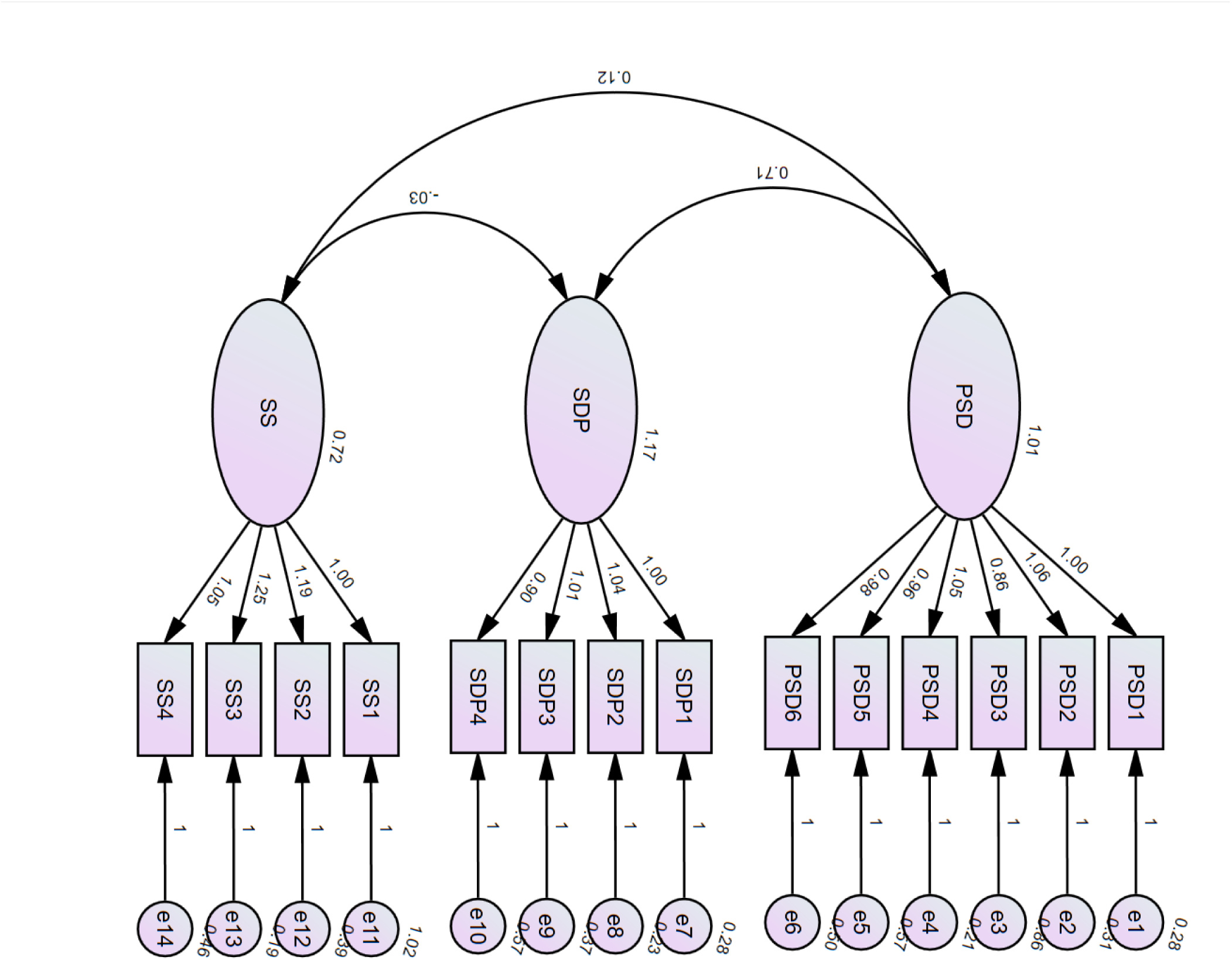
Confirmatory factor analysis of the three-factor model of the CV-HPSDS (N=1,456)

**Table 4.** The results of the model fit and its acceptance criteria (N=1456)

| Fit index | Fit standard | Modified result |
| --- | --- | --- |
| $\chi^2/\text{df}$ | <8.00 | 7.031 |
| NFI | >0.90 | 0.967 |
| GFI | >0.90 | 0.949 |
| CFI | >0.90 | 0.972 |
| IFI | >0.90 | 0.972 |
| TLI | >0.90 | 0.965 |
| RFI | >0.90 | 0.960 |
| SRMR | <0.08 | 0.044 |
| RMSEA | <0.08 | 0.064 |

### Decision to delete and re-distribution items

PSD 1 and 13 were excluded from the analysis due to their cross-factor loading on “Patient Social Discrimination” and “Social Disease Perspective”. Additionally, PSD 11 was eliminated based on the results of the CFA. The 14-item satisfies the acceptance standards of model fitting (Table 4). In response, PSD 12 was grouped with PSD 8-11 to form a new dimension called “patient social discrimination”. The dimension of “social harassment” was removed and the corresponding items were redistributed to other dimensions. Subsequently, the 14 remaining items were subjected to EFA, yielding excellent and stable psychometric parameters (KMO=0.912, Bartlett’s test χ2=18786.223, p<0.001). Notably, no items exhibited a factor loading >0.4 in other dimensions, and the cumulative explained rate was 76.563%. (Table 5)

**Table 5.**
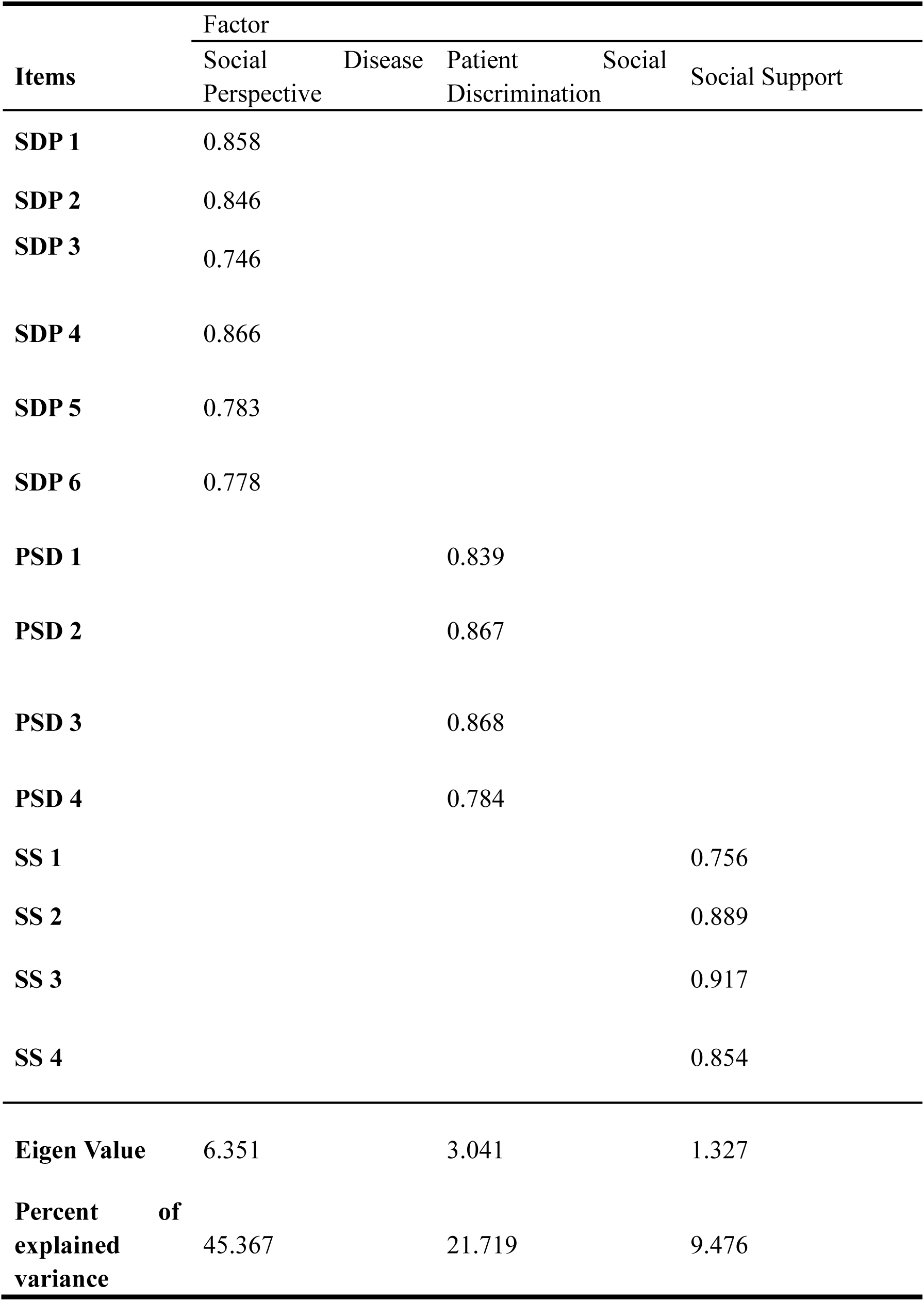
Exploratory Factor Analysis of Corrected HIV/AIDS Public Stigma and Discrimination Scale.

### Internal Consistency

The findings of this study demonstrate the stability of the scale and indicate excellent internal consistency. The total Cronbach’s coefficient of the scale was found to be 0.869, while the alpha coefficients of the subscales ranged from 0.875 to 0.927. Furthermore, the McDonald’s omega coefficient was 0.883. Taken together with the Cronbach’s alpha coefficient, these results suggest that the scale exhibits strong internal consistency.

### Known-group validity

The present study provides conclusive evidence in support of the anticipated distinction, thereby attesting to the construct validity of the scale. The findings indicated that females residing in urban areas, with university-level education and a major in medicine, reported significantly lower stigma and discrimination scores compared to other participants. Notably, although prior investigations did not specifically target female undergraduates, the current results are consistent with the findings of the preliminary study.

### Convergent validity and Discriminant validity

The present study further evaluated the construct validity of the model by conducting composite reliability and AVE testing, which yielded satisfactory results. The AVE values of the three subscales were 0.691, 0.759, and 0.658, respectively, exceeding the criterion of 0.5, indicating good convergent validity. Additionally, the CR values of the subscales ranged from 0.883 to 0.930, demonstrating that the theoretical constructs were closely related to each other and the assumptions were consistent and stable with the underlying constructs. Furthermore, divergent validity was assessed by calculating the square AVE and correlation among subscales, which showed that the square AVE, an indicator of convergence, was greater than the internal correlation among subscales. All dimensions reached a statistically significant difference (p<0.001). After analyzing the psychometric properties, we concluded that the CV-HPSDS had excellent structure and was suitable for use in the general population in China. (Table 6)

**Table 6.**
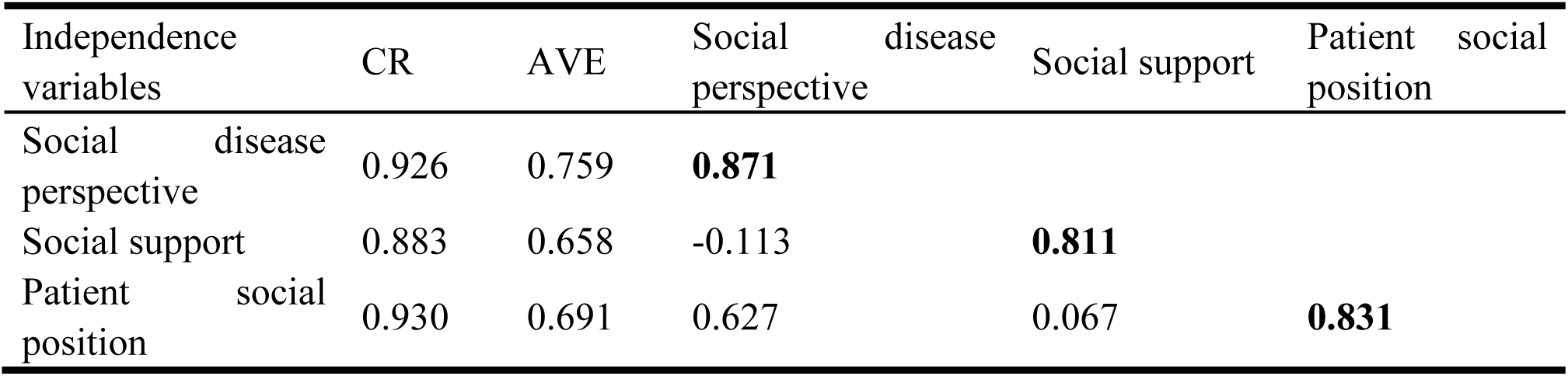
The divergent validity and convergent validity of scale.

## Discussion

The present study expounds upon the process of adapting and conducting psychometric testing on a novel HIV/AIDS stigma and discrimination scale tailored towards Chinese university students, thereby offering a significant insight into the general population’s attitudes towards stigma and discrimination. XXX As such, this information holds immense value for future researchers who aim to investigate related works in this field.

Although the factor structure and items of the CV-PHSDS were re-distributed, the scale maintained excellent internal consistency with high factor loadings and correlations. After the analysis and adaption, a 14-item adapted scale was found to contribute to stable construct validity and better divergent validity. Despite the insufficient internal consistency of the “social support” items, these items remain important in the context of stigma and discrimination and are essential for assessing public attitudes towards PLHIV in real-world settings. This content plays a crucial role in the stigma in healthy settings, social exclusion, quality of life and unfair treatment where the vital essence of PLHIV have attracted researchers to understand[22][23][24][25]. Consequently, as a critical component of stigma and discrimination, the “social support” subscale remains and assesses the public attitudes towards PLHIV in the real world. Since cross-loading items was removed from the scale, we finally obtained a short version and no cross-loading in the exploratory factor analysis through which we believed that the adapted version of the scale has better psychometric properties than the original version. The scree plot convincingly supports the adoption of the 3-factor model after adaption. The adapted version and the psychometric parameters show the stable structure of the scale, explaining 76.597% variance. Moreover, the results of internal consistency analysis were showed by Cronbach’s alpha coefficients and McDonald’s omega coefficient and test-retest reliability is 0.869, 0.883 and 0.857 respectively. And the adapted version showed better results than the original version in Iran[16]. The multidimensional approach, including composite reliability and divergent validity, supported the construct validity of the Chinese version and demonstrated that the 3 dimensions can represent typical aspects of stigma and distinguish between each other. Results indicate that the adapted version of the scale has strong psychometric properties and can be applied to Chinese university students for further research.

After expert analysis, it is recommended that certain items containing illegal behaviors that infringe on the rights of PLHIV should be omitted from the pre-final version. Due to the influence of traditional values and the strong emphasis on family-centered relationships in China, families are often unwilling to abandon their loved ones and are motivated by a sense of “love” and “filial piety” to refrain from discriminating against those with HIV/AIDS[26]. On the grounds of the cultural factors, item 8 “Families of AIDS feel humiliated to have such family member” was removed from the adapted scale to better align with cultural factors in China. Additionally, the lack of clarity in translation of certain questions from the original scale developed in Iran created a gap between language and culture, further emphasizing the importance of adapting scales to specific cultural contexts.

Numerous scales have been developed to measure the psychometric aspects of stigmas and the attitudes of the general population towards People Living with HIV (PLHIV). Although these scales have demonstrated high levels of reliability and validity, their limitations include the lack of assessment of diverse and large enough sample sizes in research studies[15][27]. To our knowledge, reliable scales for measuring HIV/AIDS-related stigma have been lacking, leading previous researchers to resort to interviews and incomplete item sets[28]. For instance, the questionnaire concerning discriminatory attitudes and knowledge was adapted and adopted by Xie H[29]. The study aimed to assess the stigmatization level of doctors and nurses in health settings where they confronted the risk of occupational exposure. Due to the professional background of doctors and nurses, it is tough to apply the scale for the general population. In comparison, the Chinese version of Zelaya’s HIV-related Stigma Scale (CVZHSS) seems to be promising measurement containing 4 dimensions of stigma[19]. The CVZHSS was translated and adapted by Xing and colleagues applied for college students and rural-urban migrants[30][31]. Based on the stigma theory[32], the scales developed by Berger et al[20] and Zelaya et al[19] integrate multidimensional constructs of stigma and discrimination. These scales employ indirect measurement items (e.g., fear of disease transmission) to assess the cognitive and behavioral origins of stigmatized attitudes.

In addition, for future development and application of the scale, we recommend including items that measure the level of support and encouragement towards PLHIV, such as should people living with HIV/AIDS be allowed to freely participate in various social activities? Such items can be useful in assessing the extent of stigma and discrimination[33][34]. As previously mentioned, the newly adapted version of the scale is beneficial in promoting work on stigma and discrimination. To our knowledge, we are the first to translate, adapt, and assess the HIV/AIDS stigma and discrimination scale in China. We also encourage the use of more diverse samples and application of the scale to the general population throughout the country.

The present study demonstrated strong reliability and validity of the newly adapted scale, providing a basis for further refinement towards a “gold standard” and the establishment of a criterion for future scales. Compared to previous researches, our scale demonstrated improved psychometric properties, with high factor loadings and internal consistency of all items exceeding 0.7. Additionally, the development of a short and valid instrument facilitates broader applications to more diverse samples in China.

In conclusion, our study successfully validated the Chinese version of the HIV/AIDS stigma and discrimination scale from a general population perspective, with high levels of reliability and validity demonstrated in terms of internal consistency, construct validity, discrimination validity, and convergent validity. The scale underwent rigorous forward-back translation and cultural adaptation to ensure its applicability in measuring stigma and discrimination towards PLHIV and AIDS patients. By introducing this new measurement tool to China, we have opened up a powerful and promising possibility for stigma research and the fight against stigma and discrimination.

## limitation

Regarding the translation and adaptation process of our study, we also acknowledge some limitations. Firstly, the online convenience sampling method used in our study may have some shortcomings, such as participants unconsciously filling in the blanks or ignoring/misreading important information without proper guidance. Additionally, our study did not include items related to fear or knowledge about HIV transmission, which are important factors included in other stigma scales. We believe that including these items in the CV-HPSDS can improve its comprehensiveness and usefulness. Furthermore, the generalizability of the findings should be interpreted with caution, as the data were collected from a specific university student population that systematically differs from the general population in terms of age distribution, educational background, and social experience. Future studies could validate the results’ applicability through expanded sampling strategies. Lastly, the failure to obtain ethics approval has restricted the publication of research findings in academic journals and their implementation in practical applications, thereby hindering the dissemination of the research outcomes and diminishing their real-world value.

## Author Contributions

Xiaohan Wang: formal analysis, methodology, project administration, supervision, validation.

Zhengyang Pan: formal analysis, methodology, resources, validation, visualization.

Jingjing Zhao: Conceptualization, data curation, formal analysis, writing – original draft, writing – review & editing.

Runqing Liu: Conceptualization, data curation, formal analysis, writing – original draft, writing – review & editing.

Zhe Wu: formal analysis, investigation, writing – original draft, writing – review & editing.

Xiang Chen: formal analysis, investigation, resources, writing – original draft.

## Acknowledgments

We gratefully acknowledge Professor Mahlagha Dehghan for granting us permission to use the original version of the HIV/AIDS Stigma and Discrimination Scale, which provided a solid foundation for this study. We also extend our sincere appreciation to all healthcare professionals and experts—including Chunsheng Ding, Shaojun Xu, Yuan Tian, Ning Zhang, and others—who responded to our invitation and participated in the translation and adaptation process. Our special thanks go to all questionnaire participants for their valuable contributions. We affirm that all personal information will be kept strictly confidential, and data will be used solely for academic research purposes.

## Data Availability Statement

The adapted process and data can be found at https://osf.io/8au7y/, further questions can email to the corresponding.

## Ethics Statement

This study did not involve animal or human clinical trials and was conducted as a psychometric investigation. In accordance with the ethical principles outlined in the Declaration of Helsinki, informed consent was obtained from all participants prior to their involvement. The anonymity and confidentiality of participants were ensured, and participation was entirely voluntary.

## References

[1] UNAIDS. (2024). Fact sheet - Latest global and regional statistics on the status of the AIDS epidemic. UNAIDS. https://www.unaids.org/en/resources/documents/2024/UNAIDS_FactSheet.

[2] Zhang, J., Ma, B., Han, X., Ding, S., & Li, Y. (2022). Global, regional, and national burdens of HIV and other sexually transmitted infections in adolescents and young adults aged 10-24 years from 1990 to 2019: a trend analysis based on the Global Burden of Disease Study 2019. The Lancet. Child & adolescent health, 6(11), 763 – 776. 10.1016/S2352-4642(22)00219-X.

[3] Wong, C. S., Wei, L., & Kim, Y. S. (2023). HIV Late Presenters in Asia: Management and Public Health Challenges. AIDS research and treatment, 2023 9488051. 10.1155/2023/9488051.

[4] Teshale, A. B., & Tesema, G. A. (2022). Discriminatory attitude towards people living with HIV/AIDS and its associated factors among adult population in 15 sub-Saharan African nations. PloS one, 17(2), e0261978. 10.1371/journal.pone.0261978.

[5] Aziz, M. M., Abdelrheem, S. S., & Mohammed, H. M. (2023). Stigma and discrimination against people living with HIV by health care providers in Egypt. BMC health services research, 23(1), 663. 10.1186/s12913-023-09676-1.

[6] UNAIDS. (2021, June 11). High expectations | UN adopts new political declaration on HIV/AIDS [Web page]. Joint United Nations Programme on HIV/AIDS China Website. https://www.aids.org.cn/cn/index/7669_2307168_61820.

[7] Asrina, A., Ikhtiar, M., Idris, F. P., Adam, A., & Alim, A. (2023). Community stigma and discrimination against the incidence of HIV and AIDS. Journal of medicine and life, 16(9), 1327–1334. 10.25122/jml-2023-0171.

[8] Hu, Y., Zhou, X., Fan, X., Bi, R., Deng, Y., Li, H., Peng, X., Luo, D., Zhao, H., Guo, Z., He, L., & Zou, H. (2025). Prevalence, correlates and solutions to people with HIV in China being refused treatment for diseases not related to HIV: a mixed-methods study. Journal of the International AIDS Society, 28(4), e26443. 10.1002/jia2.26443.

[9] Thornicroft, G., Sunkel, C., Alikhon Aliev, A., Baker, S., Brohan, E., El Chammay, R., Davies, K., Demissie, M., Duncan, J., Fekadu, W., Gronholm, P. C., Guerrero, Z., Gurung, D., Habtamu, K., Hanlon, C., Heim, E., Henderson, C., Hijazi, Z., Hoffman, C., Hosny, N., … Winkler, P. (2022). The Lancet Commission on ending stigma and discrimination in mental health. Lancet (London, England), 400(10361), 1438–1480. 10.1016/S0140-6736(22)01470-2

[10] Letsela, L., Jana, M., Pursell-Gotz, R., Kodisang, P., & Weiner, R. (2021). The role and effectiveness of School-based Extra-Curricular Interventions on children’s health and HIV related behaviour: the case study of Soul Buddyz Clubs Programme in South Africa. BMC public health, 21(1), 2259. 10.1186/s12889-021-12281-8.

[11] Kimera, E., Alanyo, L.G., Pauline, I. et al. Community-based interventions against HIV-related stigma: a systematic review of evidence in Sub-Saharan Africa. Syst Rev 14, 8 (2025). 10.1186/s13643-024-02751-6.

[12] Fauk, N. K., Hawke, K., Mwanri, L., & Ward, P. R. (2021). Stigma and Discrimination towards People Living with HIV in the Context of Families, Communities, and Healthcare Settings: A Qualitative Study in Indonesia. International journal of environmental research and public health, 18(10), 5424. 10.3390/ijerph18105424.

[13] Visser, M. J., Kershaw, T., Makin, J. D., & Forsyth, B. W. (2008). Development of parallel scales to measure HIV-related stigma. AIDS and behavior, 12(5), 759 – 771. 10.1007/s10461-008-9363-7.

[14] López, A., Rafful, C., Orozco, R., Contreras-Valdez, J. A., Jiménez-Rivagorza, L., & Morales, M. (2023). HIV Stigma Mechanisms Scale: Factor Structure, Reliability, and Validity in Mexican Adults. AIDS and behavior, 27(4), 1321–1328. 10.1007/s10461-022-03868-2.

[15] Huang, F., Chen, W. T., Shiu, C. S., Lin, S. H., Tun, M. S., Nwe, T. W., Oo, Y. T. N., & Oo, H. N. (2021). Adaptation and validation of a culturally adapted HIV stigma scale in Myanmar. BMC public health, 21(1), 1663. 10.1186/s12889-021-11685-w.

[16] Mokhtarabadi, S., Sharifi, H., Rad, A. A. R., Iranpour, A., & Dehghan, M. (2020). Development and Validation of HIV/AIDS Stigma and Discrimination Scale in Southeast Iran: The General Population Viewpoint. Journal of the International Association of Providers of AIDS Care, 19, 2325958220963601. 10.1177/2325958220963601.

[17] Kubátová, A., Fialová, A., Stupka, J., Malý, M., Hamplová, L., & Sedláčková, S. (2023). Stigmatization and discrimination of people living with HIV in the Czech Republic: a pilot study. Central European journal of public health, 31(3), 210 – 216. 10.21101/cejph.a7782.

[18] Jones, P. S., Lee, J. W., Phillips, L. R., Zhang, X. E., & Jaceldo, K. B. (2001). An adaptation of Brislin’s translation model for cross-cultural research. Nursing research, 50(5), 300–304. 10.1097/00006199-200109000-00008.

[19] Zelaya, C. E., Sivaram, S., Johnson, S. C., Srikrishnan, A. K., Solomon, S., & Celentano, D. D. (2008). HIV/AIDS stigma: reliability and validity of a new measurement instrument in Chennai, India. AIDS and behavior, 12(5), 781 – 788. 10.1007/s10461-007-9331-7.

[20] Berger, B. E., Ferrans, C. E., & Lashley, F. R. (2001). Measuring stigma in people with HIV: psychometric assessment of the HIV stigma scale. Research in nursing & health, 24(6), 518–529. 10.1002/nur.10011.

[21] Goffman, & Erving. (1963). Stigma: notes on the management of spoiled identity. American Journal of Sociology.

[22] Adhiambo, H. F., Ngayo, M., & Kwena, Z. (2022). Preferences for accessing sexual and reproductive health services among adolescents and young adults living with HIV/AIDs in Western Kenya: A qualitative study. PloS one, 17(11), e0277467. 10.1371/journal.pone.0277467.

[23] Marziali, M. E., McLinden, T., Card, K. G., Closson, K., Wang, L., Trigg, J., Salters, K., Lima, V. D., Parashar, S., & Hogg, R. S. (2021). Social Isolation and Mortality Among People Living with HIV in British Columbia, Canada. AIDS and behavior, 25(2), 377 – 388. 10.1007/s10461-020-03000-2.

[24] Rayanakorn, A., Ong-Artborirak, P., Ademi, Z., & Chariyalertsak, S. (2022). Predictors of Stigma and Health-Related Quality of Life Among People Living with HIV in Northern Thailand. AIDS patient care and STDs, 36(5), 186–193. 10.1089/apc.2022.0035.

[25] Penn, T. M., Trost, Z., Parker, R., Wagner, W. P., Owens, M. A., Gonzalez, C. E., White, D. M., Merlin, J. S., & Goodin, B. R. (2019). Social support buffers the negative influence of perceived injustice on pain interference in people living with HIV and chronic pain. Pain reports, 4(2), e710. 10.1097/PR9.0000000000000710.

[26] Liu CM, Shang, Y. M., Jiao, C. H., Sun, H. Y. (2014). On the dignity and rights of the death: Take cancer patients’ suicide as an example [In Chinese]. Medicine & Philosophy. (A), 35 (03), 20–22.

[27] Luz, P. M., Torres, T. S., Almeida-Brasil, C. C., Marins, L. M. S., Bezerra, D. R. B., Veloso, V. G., Grinsztejn, B., Harel, D., & Thombs, B. D. (2020). Translation and validation of the Short HIV Stigma scale in Brazilian Portuguese. Health and quality of life outcomes, 18(1), 322. 10.1186/s12955-020-01571-1.

[28] Alshouibi, E., & Alaqil, F. (2019). HIV-Related Discrimination among Senior Dental Students in Jeddah. Journal of International Society of Preventive & Community Dentistry, 9(3), 219–224. 10.4103/jispcd.JISPCD_420_18.

[29] Xie, H., Yu, H., Watson, R., Wen, J., Xiao, L., Yan, M., & Chen, Y. (2019). Cross-Cultural Validation of the Health Care Provider HIV/AIDS Stigma Scale (HPASS) in China. AIDS and behavior, 23(4), 1048–1056. 10.1007/s10461-018-2312-1.

[30] Xing, H., Yu, W., & Li, Y. (2016). Measuring and assessing HIV/AIDS stigma and discrimination among migrant workers in Zhejiang, China. BMC public health, 16, 845. 10.1186/s12889-016-3518-7.

[31] Ruan, F., Fu, G., Zhou, M., Luo, L., Chen, J., Hua, W., Li, X., Chen, Y., Xia, X., Xiong, Y., Chen, Y., Shi, B., Lu, S., Zhang, H., Wu, D., Liu, Y., Zhan, J., & Wang, J. (2019). Application of the Chinese version of Zelaya’s HIV-related stigma scale to undergraduates in mainland China. BMC public health, 19(1), 1708. 10.1186/s12889-019-8054-9.

[32] Link, B. G., & Phelan, J. C. (2006). Stigma and its public health implications. Lancet (London, England), 367(9509), 528–529. 10.1016/S0140-6736(06)68184-1.

[33] Deacon, H. (2006). Towards a Sustainable Theory of Health-Related Stigma: Lessons from the HIV/AIDS Literature. Journal of Community & Applied Social Psychology, 16(6), 418–425. 10.1002/casp.900.

[34] Corrigan, P. W. (2000). Mental health stigma as social attribution: Implications for research methods and attitude change. Clinical Psychology: Science and Practice, 7(1), 48 – 67. 10.1093/clipsy.7.1.48.

